# Post Operative Pain Management (POPM) and Patient Satisfection : A Tertiary Care Hospital Experience

**DOI:** 10.1101/2021.05.11.21257045

**Authors:** Jannatul Ferdoush, Rajib Pal Chowdhury, Fatema Johora, Fatiha Tasmin Jeenia, Maliha Ata, Kohinoor Parveen, Sharif Mohammad Towfiq Hossain, Shagorika Sharmeen, MD Sayedur Rahman

## Abstract

**Background:** Pain is an inevitable part of the postoperative experience. Current study was designed to assess the pattern of analgesic use, the adequacy of post operative pain control by documenting pain scores, and patient satisfaction by documenting the pain perception score of the postoperative patients 48 hours after surgery.

**Materials and methods:** This was a formative research and includes a mix of descriptive cross-sectional study carried out in the department of Gynecology and Obstetrics, General surgery, and Orthopedics surgery of Chattagram Maa -O- Shishu Hospital Medical College (CMOSHMC) during the month of January, 2018 to July, 2019.

**Result:** A total of 180 patients undergoing surgery were included in this study. Baseline data were collected both retrospectively and prospectively. Only analgesic used immediately after operation in three departments were Pethidine (100%). Diclofenac sodium suppository were used in appendectomy and cesarean section (50/60, 83%) and (26/60, 43%) respectively. Mostly, Diclofenac Sodium IV (55/60, 91%) was used in lower limb surgery. The maximum pain score were 5.3 (*SD*=2.3), 5.6 (*SD* =1.9), and 6.1 (*SD* =1.3) during coughing in the first 8 hour and minimum pain score 2.4 (*SD* =1.0), 2.2 (*SD* = 0.8) and 1.7 (*SD*=1.3) at rest 48 hours after cesarean section, appendectomy and lower limb surgery respectively. But pain score eventually increased during coughing 48 hours after surgery. After 48 hours of surgery maximum pain perception score 3.9 (*SD* = 0.9) in lower limb surgery and minimum perception score were 3.5 (*SD* =1.8) in cesarean section. Majority of the appendectomy patient (76%) satisfied with pain management where as only 43% satisfied in case of cesarean section.

**Conclusion:** This study enabled the researchers to get a precise picture of the current state of POPM in different hospitals, as well as identify particular needs for improving such practices in health care facilities and implementing an educational intervention in order to improve the post operative pain management.

## Introduction

Postoperative pain is a form of acute pain that is often accompanied by a neuroendocrine stress response that is proportional to the intensity of pain. After surgery, about 80% of patients reported severe pain [1]. Suboptimal pain management can result in postsurgical consequences such as deep vein thrombosis, pulmonary embolus, and pneumonia, which have a negative impact on patient safety, hospital performance, and the cost of treatment [2]. According to evidence, the adequacy of counseling received, nature of treatment, preoperative knowledge, actual experience of pain, and overall pain experience are factors correlated with pain management satisfaction [3].

Freedom from pain is necessary for patient. For many years, Patients with abdominal pain were not allowed enough analgesia as it masked the sign required for diagnosis [4]. That was incorrect. When you have the fact, it’s much easier to debunk myths.

The earliest anesthetic was provided for painless surgery in 1986 [5]. Some 150 years later, patients should not have had to suffer unrelieved pain somewhere in the hospital. Pain is considered as the “fifth vital sign,” [6] and it can be measured bedside during routine postoperative care and reported at frequent interval so that all members of the health care stuff have a better view of the pain.

The major concern in surgical procedures is post-operative pain. Repeated pain scoring in the postoperative period is an integral part of lowering the incidence and severity of acute postoperative pain, as well as improving patient comfort and satisfaction [7].

The most intense post-operative pain occurs within the first 24 hours after surgery and normally subsides within 48 hours [8]. According to many sources, the patient’s own verbal report [9] and the use of a pain scale that can establish a shared language between patients and healthcare providers is the simplest and most reliable index of pain [10].

Along with a pain score, a satisfaction score should be obtained. This blend will help to ensure that unattended, needless pain is not overlooked. A successful pain management program relies on responsive analgesia control with good patient communication. As a result, national and international recommendations emphasize the importance of conducting pain assessment at rest and during activity on a regular basis. [11-13].

There are many flaws with current post operative pain management such as physician’s knowledge, patient records, pain evaluation, and recommendations. On the basis of doctors’ scientific medical expertise and personal understanding, clinical decisions are made, but there is also a substantial disparity in what they know and what they do. Clinicians, on the other hand, often follow the guidelines to a certain extent and often depend on their own professional expertise [14]. Although practice pattern seems to be changing, initiatives should be stepped up to accelerate the transition to a pain-free health facility [15].

With renewed emphasis on pain relief services and the adoption of new pain management standards, many patients experience significant pain following surgery. Difficulties in assessing pain, inconsistency in medication and dosing, and a lack of expertise on the part of caregivers may all be the obstacles. Specific pain relief needs can vary greatly between patients, and an individual pain assessment is needed to capture these differences [16].

Since there is a lack of data in our country that provides adequate and detailed information about POPM, this study was designed to assess the pattern of analgesic use, the adequacy of post operative pain control by documenting pain scores, and patient satisfaction by documenting the pain perception score of the postoperative patient after 48 hours of surgery. It can be presumed that the findings of our study could identify the specific needs to strengthen such practices in healthcare facilities by identifying areas of the pain control process that fall short of minimum standards.

## Materials and Methods

This was a formative research and includes a mix of descriptive cross-sectional study carried out in the Gynecology and Obstetrics, General surgery and Orthopedics department of Chattagram Maa -O- Shishu Hospital Medical College (CMOSHMC) during June 2018 to July, 2019. Baseline data were collected both retrospective and prospectively. For collection of retrospective data, treatment records of inpatients 30 cases of surgery from each department were selected by random sampling qualifying every odd number registered patients from the total cases of previous six months. Pattern of analgesic used were sorted out from these patients. Patient satisfaction for these retrospective patients could not be evaluated. For collecting prospective data/current data, treatment sheets of the 30 cases of admitted inpatients from each department available on nurse’s desk were selected randomly. The surgical wards chosen for this study were those representing 80% of inpatients receiving postoperative analgesia, i.e., orthopedics, gynecological and general surgery wards. In patients with multiple surgical operations, patients suffering from Diabetes mellitus, Hypertension, Bronchial asthma, Renal disease, any Cardiac diseases were excluded from the study. If patient received analgesic for any reason within 24 hours of surgery and incidence of any infection were excluded also.

### Procedure of Data collection

For retrospective portion of the study, baseline data was collected from treatment record of selected hospital. Treatment sheets of previous post operative patients were included on the basis of inclusion and exclusion criteria. Then prospective study was carried out in the departments of Gynecology and Obstetrics, General surgery, Orthopedics surgery regarding pain assessment and documentation of pain score in the patient chart.

### Postoperative pain assessment

Before surgery, patient was briefly counseled regarding pain assessment technique in an understandable language that is suitable for the patient. Postoperative pain assessment was performed with interviews, when patients were asked about the presence of pain at the moment of such assessment. Assessment was done at 0 hours, 2 hours, 8 hours, 12 hours, 24 hours, 36 hours and 48 hours at rest and during movement after intervention and pain score was documented in patient chart. Perception score of patient satisfaction regarding pain management were documented after 24 and 48 hours interval. Aiming to quantify this symptom, a Visual analogue scale was used [17].

## Result

The Study enrolled a total of 180 patients. 30 cases of appendectomy, 30 cesarean section, and 30 lower limb surgeries were surveyed from previously undergone surgery record files, and prospectively 30 appendectomy, 30 cesarean section, and 30 lower limb surgery patients were included in this study.

The mean patient age was 44.2 years, and 32.3% were female and 67.7% were male (Table not shown). Quality of record keeping was satisfactory as there was no significant difference between prospective and retrospective data.

Table I showed that, the only analgesic used immediate after operation in three departments were pethidine (100%). After 6 hours of surgery, Diclofenac sodium suppository 8 hourly were used in appendectomy and cesarean section (26/60, 43%) and (50/60, 83%) respectively. Mostly, Diclofenac Sodium IV (55/60, 91%) 8 hourly was used in lower limb surgery after 6 hours of surgery. The prescribed oral post operative analgesic was Paracetamol alone (70%) or in combination with Tramadol (31%) mostly in cesarean section.

**Table I:** Pattern of Analgesics during the First 48 Hours of Surgery.

|  | Abdominal surgery<br>(Appendectomy)<br>N=60 | Obstetric surgery<br>(Cesarean section)<br>N=60 | Orthopedic surgery<br>(Lower limb surgery)<br>N=60 |
| --- | --- | --- | --- |
| Pethidine IM (Immediate after surgery) | 60(100%) | 60(100%) | 60(100%) |
| Diclofenac sodium suppository (8hourly) | 26(43%) | 50(83%) | 23(38%) |
| Diclofenac sodium IV(12 hourly) | 0(0%) | 0(0%) | 55(91%) |
| Ketorolac IV(12 hourly) | 10(16%) | 0(0%) | 5(9%) |
| Tramadol suppository(8 hourly) | 3(5%) | 10(16%) | 24(40%) |
| Paracetamol oral if needed | 12(20%) | 42(70%) | 5(8%) |
| Paracetamol plus Tramadol oral if needed | 0(0%) | 19(31%) | 7(12%) |

Table II showed that, the intensity of pain on arrival in the recovery room (immediate postoperative period) was 0±0 for all post operative patient. The maximum pain score 8 hours after cesarean section were 3.4±1.9 and 5.3 ±2.3 at rest and coughing respectively. Pain score was decreased 2.4±1.0 during rest but increased during coughing were 3.4±1.3 at 48 hours after cesarean section. In case of appendectomy maximum pain score 2 hours after operation were 4.4±2.3 and 5.6 ±1.9 at rest and coughing respectively and minimum pain score was 2.2±0.8 and 4.3±0.7 at rest and coughing respectively 48 hours after surgery. In lower limb surgery, maximum pain score 2 hours after operation 5.4±0.9 and 6.1±1.3 at rest and coughing respectively and minimum pain score 1.7±1.3 at rest at 48 hours after surgery. But pain score increased during coughing was 3.8±0.7 at 48 hours after surgery.

**Table II:** Post Operative Pain Scores at Different Time Interval.

| **Post Operative Pain Score** |  |  |  |  |  |  |
| --- | --- | --- | --- | --- | --- | --- |
| Expressed as (Mean ± SD) | | | | | | |
|  | **Cesarean section**<br>(N=30) |  | **Appendectomy**<br>(N=30) |  | **Lower limb surgery**<br>(N=30) |  |
| Time interval | at rest | at coughing | at rest | at coughing | at rest | at coughing |
| Immediate | 0±0 | 0±0 | 0±0 | 0±0 | 0±0 | 0±0 |
| 2 hours | 1.3±1.9 | 2.6±2.3 | 4.4±2.3 | 5.6±1.9 | 5.4±0.9 | 6.1±1.3 |
| 8 hours | 3.4±1.9 | 5.3±2 | 3.9±1.8 | 4.8 ±1.2 | 4.4±1.9 | 5.1±2.0 |
| 12 hours | 3.1±1.5 | 4.2±1.9 | 3.3±2.1 | 4.8 ±1.9 | 2.8±1.5 | 3.9±1.5 |
| 18 hours | 2.8±1.7 | 4.0±1.8 | 3.2 ±1.3 | 4.9±0.9 | 3.3±1.3 | 4.5±0.9 |
| 24 hours | 2.6±1.5 | 3.7±1.8 | 2.2±1.6 | 4.1±1.1 | 2.1±1.4 | 3.7±1.8 |
| 36 hours | 2.1±1.0 | 3.1±1.6 | 2.8±1.9 | 4.8±1.2 | 2.4±1.0 | 3.6±1.5 |
| 48 hours | 2.4±1.0 | 3.4±1.3 | 2.2±0.8 | 4.3±0.7 | 1.7±1.3 | 3.8±0.7 |

Table III showed that, maximum pain perception score 3.9±0.9 at 48 hours after lower limb surgery and minimum pain perception score 3.5±1.8 at 48 hours after Cesarean section. Majority (76%) of the Patient satisfied with pain management 48 hours after appendectomy. Whereas in case of caesarean section 43% patient mentioned satisfied with pain management after 48 hours.(fig I)

**Table III:**
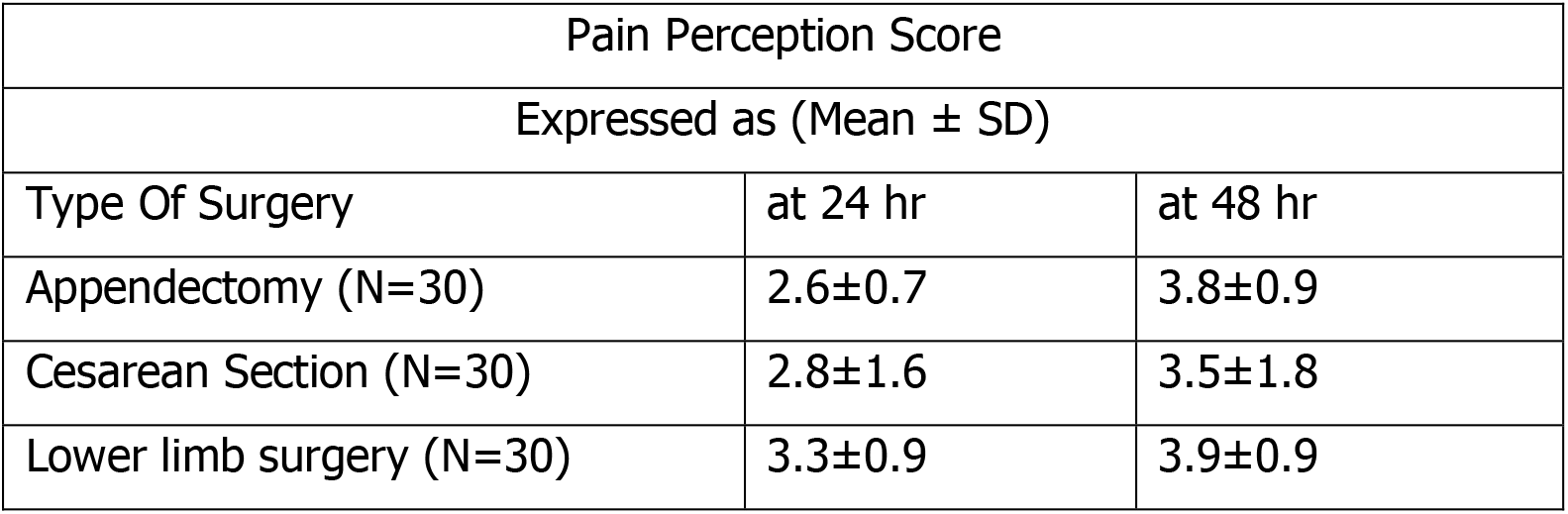
Pain Perception Scores of Patients after Surgery.

**Figure I:**
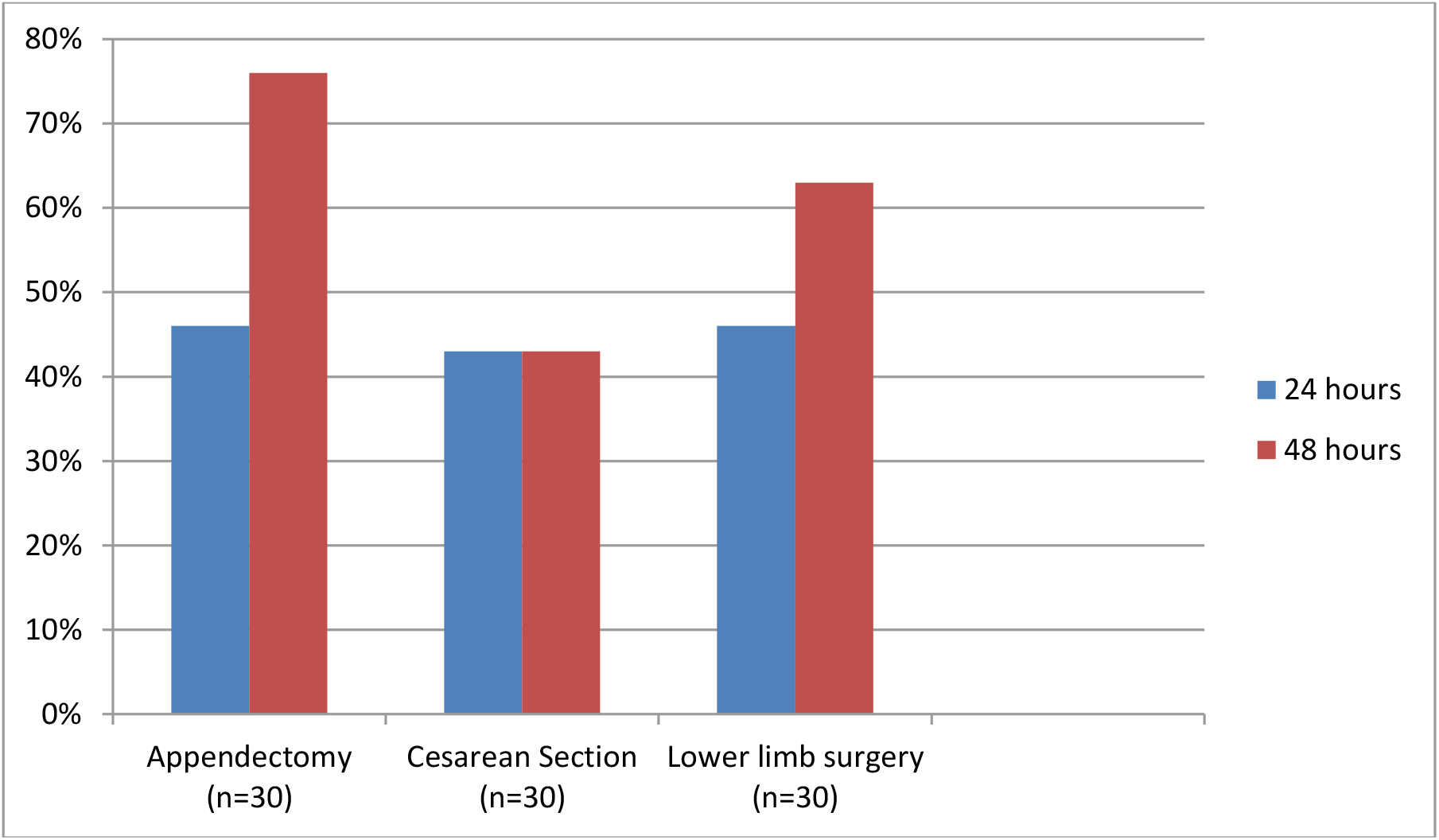
Patient Satisfaction With Pain Management.

## Discussion

This study was designed to assess the pattern of analgesic use, adequacy of post operative pain control by documenting pain score and satisfaction with post operative pain management by documenting pain perception score at 24 and 48 hours after surgery. Current study revealed that Pethidine was used immediately after operation in three departments in all cases (100%) for provision of analgesia. After 8 hours of surgery, Diclofenac Sodium suppository 8 hourly were used in appendectomy and cesarean section (26/60, 43%) and (50/60, 83%) respectively. Mostly, Diclofenac Sodium IV (55/60, 91%) 8 hourly was used in lower limb surgery after 6 hour of surgery. The prescribed oral post operative analgesic was Paracetamol alone (70%) or in combination with Tramadol (31%) mostly used in cesarean section. Frequent use of Pethidine in the day of surgery was observed in other studies conducted in Bangladesh [18-20] as efficacy of opioids were proved in the acute treatment of moderate-to-severe pain in the early postoperative period [12].

In the current study, the intensity of pain immediate postoperative period was 0±0 for all post operative patients. The maximum pain score 8 hours after cesarean section were 3.4±1.9 and ±2.3 at rest and coughing respectively. Pain score was decreased 2.4±1.0 during rest but increased during coughing were 3.4±1.3 at 48 hours after cesarean section. Since lactating mothers must stay alert and active in order to care for and breastfeed their newborns, pain was higher throughout movement. This finding is consistent with jawaid et al. (2010) study where postoperative mean rest and dynamic pain score were 3.85 ± 2.45 and 5.32 ± 2.61 at 12 hours and 2.84 ± 1.86 and 4.65 ± 2.47 at 48 hours interval [21].

In case of appendectomy maximum pain score 2 hours after operation were 4.4±2.3 and 5.6 ±1.9 at rest and coughing respectively and minimum pain score was 2.2±0.8 and 4.3±0.7 at rest and coughing respectively 48 hours after surgery. In lower limb surgery, maximum pain score 2 hours after operation 5.4±0.9 and 6.1±1.3 at rest and coughing respectively and minimum pain score 1.7±1.3 at rest 48 hours after surgery. But pain score increased during coughing was 3.8±0.7 at 48 hours after surgery. Consistent to this finding, a study in Malaysia (2018) found 95.4% of patients reported postoperative pain during the first 24 hour after surgery with the mean pain intensity score 4.13± 2.23 [22].

However, pain scores are linked with satisfaction with pain management. As a result, the patient should be reassessed to see whether he or she is satisfied with the management. It has been consistently shown that medical personnel often misinterpret the severity of pain that patients are feeling; resulting in analgesic administration is either delayed or inadequate [10]. Our study revealed that, maximum perception score 3.9±0.9 at 48 hours after lower limb surgery and minimum perception score 3.5±1.8 at 48 hours after cesarean section. Majority (76%) of the Patient satisfied with pain management after 48 hours of appendectomy whereas in case of caesarean section 43% of patients satisfied with pain management after 48 hours. According to Chung JW (2003) study, 66% patients were satisfied with their post-operative pain control [23].

Patients’ involvement in their pain management may raise pain awareness, but it greatly improves their satisfaction with postoperative pain control [23]. This result is in line with the findings of other research, which found that the majority of patients were satisfied with their post-operative pain management [21, 24]. These findings can be explained by the fact that patients have generally assumed that postoperative pain is unavoidable and that they must suffer through it, and that only a small percentage of patients were aware of the standard of health care they could receive and the possible benefits of satisfactory pain relief [23, 25].

Limitation of current study is that the pain severity was not correlated with other variables such as sex, education, method of anesthesia, or prior surgical history in our research. This study did not made any association between pain intensity score and patient satisfaction with overall pain management.

## Conclusion

Postoperative pain management should be prioritized as an integral component of surgical patient care. Using pain intensity as an indicator would facilitate healthcare providers to monitor the physical recovery of the majority of patients in routine clinical practice. Current study contributes to the perceptive of patients’ experiences on pain assessments, which was previously inadequate. Routine pain scoring in the postoperative period is a key component in minimizing the incidence and severity of acute postoperative pain, as well as improving patient safety and satisfaction.

## Data Availability

This was a formative research and includes a mix of descriptive cross-sectional study carried out in the department of Gynecology and Obstetrics, General surgery, and Orthopedics surgery of Chattagram Maa -O- Shishu Hospital Medical College (CMOSHMC) during the month of January, 2018 to July, 2019.

